# The Study of Acute Myocardial Infarctions Triggers in North-Western Iran

**DOI:** 10.1101/2020.07.03.20145987

**Authors:** Kamal Khademvatani, Amin Sedokani, Sima Masudi, Parisa Nejati, Mir Hosein Seyyed Mohammadzad, Alireza Rahmani

## Abstract

**Background:** Myocardial infarction (MI) is one of the important cardiovascular diseases. A trigger is an external stimulus, potential to create a pathological change leading to a clinical event. In addition to the risk factors of myocardial infarction, MI triggers play critical roles in the incidence of acute MI.

**Methods and Results:** This is a cross-sectional study of patients with the first acute myocardial infarction in Seyedoshohada heart center of Urmia, Iran. 254 patients were enrolled in the study within one year of study. After 48^h^ of hospitalization, the patients were provided with the questionnaire to collecting the history of the disease ad triggers. Out of 220 (86.4%) patients with STEMI and 34 (13.6%) patients with NSTEMI, there were significant differences (p <0.05) in AMI triggers with LVEF (0.03), gender (0.027), residency and living area (0.039), occupation (0.002), smoking (0.008), abnormal serum TG levels (0.018) and the season of AMI occurrence (0.013). The mean age for AMI patients was 60.4±12.97 years old with a mean BMI of 26.65±4.35 kg/m^2^.

**Conclusion:** In addition to classic risk factors of myocardial infarction, health care systems must pay more attention to triggers that may induce an acute myocardial infarction in people with predisposing factors especially in the male sex, stressful and hand working jobs, and psychological and mental tension patients.

## Introduction

Over the last decade, cardiovascular disease (CVD) has become the first cause of death across the world [1, 2]. Myocardial infarction (MI) is one of the most important cardiovascular diseases. Myocardial infarction is recognized by clinical features, a rise of serum necrosis biomarkers of cardiac myocytes, and by aging, or pathological studies [3]. Today, the main pathophysiology of acute myocardial infarction (AMI) is almost well-known to all and the importance of chronic process leading to myocardial infarction is known and there is a wide agreement on it, but there is another basic question about the relationship between some acute stresses, activities or circumstances (called triggers) and AMI in the patients with predisposing factors. By the way, some studies have demonstrated that increase hospital admission after acute emotional stresses, such as natural and industrial disasters and terroristic attacks [4].

A trigger is an external stimulus, potential to create a pathological change leading to a clinical event [5]. In addition to the effects of long-term stressors such as prolonged exposure to high levels of air pollution, many studies have shown that there is a significant increase in the risk of cardiovascular events immediately after behavioral, environmental, and psychological stimuli [4, 6-8]. After studies demonstrated an increase in the incidence of myocardial infarction after the Athens earthquake, lots of studies were done on this case such as; circadian variation and onset of AMI [9], emotional triggering [10] relationship between cocaine use and immediate AMI [11, 12], or the relationship between anger and acute coronary events [13].

According to the prevalence of cardiovascular and coronary heart disease in mortality, morbidity, and cost to health systems, with studying of stimulators that mostly are depended to lifestyles and cultures, surely with declaring of triggers leading to CHDs with extended epidemiological studies, health systems can increase public knowledge of the risk and by adding preventive and therapeutic plans to the patient treatment processes, can significantly decrease the incidence of a cardiovascular event and improve health services.

## Material and Methods

A total number of 254 patients with acute myocardial infarction were enrolled in the study. The enrolled patients were all of the patients with AMI referred to Seyedoshohada heart center of West Azarbayjan of Iran through one year. After the approval of the study proposal in the research and ethics committee of Urmia medical sciences university and obtaining an ethics code for the designed questionnaire, the study began. The enrolling process was performed on patients admitted to the cardiac care units (CCU) and 48h was pasted after their hospitalization with a proven diagnosis of acute myocardial infarction with clinical, paraclinical and ECG specific signs and symptoms. After ensuring the proper treatment of patients, the questionnaire was used to collect information from patients. In this study, patients who were not able to respond due to the severity of the disease or cognitive impairment, as well as patients who were re-hospitalized after discharge or a sign or history of previous MI in the ECG (Q waves or static ST elevation), echocardiography, nuclear imaging or medical profile, were also excluded from the study.

### Paraclinical data

For assessment of hypertension, a history of previously known disease and non-invasive pressure monitoring used in CCU. Diagnosis of diabetes mellitus (DM) also, was assessed by the history of previous disease, classic signs (polyuria, polydipsia, and polyphagia) and paraclinical measuring of HbA1C of ≥6.4%, FBS of ≥126mg/dL and/or random BS of ≥200mg/dL.

In the study, we defined hypercholesterolemia as total cholesterol of >200mg/dL, low serum HDL levels as HDL of <50mg/dL for women and <40mg/dL for men. For serum triglycerides, we regrouped patients as with normal TG and with abnormal levels of serum TG with a cutoff point of <150mg/dL for normal and ≥150mg/dL for borderline and high levels of TG (abnormal levels of serum TG). Also, we determined LDL normal cutoff as below 130. For levels equal to or higher than 130, we placed them in a high LDL group.

### Triggers

At first, we divided triggers into 7 groups including; ‘Heavy Activity, Receiving Bad News, Overdosing of Drugs or Opiates, Insomnia or Mental tensions, Argue, Anger or Fight, Noisy Party or Overeating, and No triggers’ and we reported the descriptive analysis in the results. But due to the small size of each group, we regrouped them as Physical triggers, Mental triggers, and No triggers for final analysis.

### Ethical Statement

All of the stages of the study were under supervision and confirmation of the ethical committee of Seyedoshohada heart center and *<u>Urmia University of Medical Sciences</u>* (certificated ethical committee of Institutional Review Board). After all the patients were provided with written and oral explanations, they received written consent (informed written consent was given prior to the inclusion of subjects in the study). All patients had the opportunity to withdraw from the study at any time. Patient information remained confidential. There was not any intervention or extra cost to patients. The study has met the principles outlined in the Declaration of Helsinki.

### Data Analysis

The reported values are mean ± standard deviation and P-value are estimated based on one-way ANOVA. The reported values for other variables in the table are number (%), and P-values are estimated based on chi-square test.

## Results

Due to the small number of patients in subgroups, we separated all 254 patients into 3 groups of anterior and inferior ST elevated MI and non-ST elevated MI (Fig 1). Fig 2 shows the distribution of monthly and seasonal AMI. The demographical, clinical and paraclinical data are presented in table 1. Distribution of last 24 hours triggers, revealed that almost half of patients (53.6%) had no activity of known types. In another hand, 46.4% of patients had a mental or physical trigger for AMI. The incidence of AMI was 3 times bigger in men.

**Figure 1.**
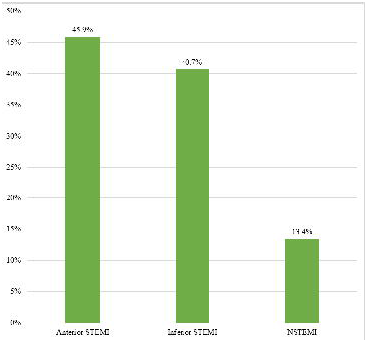
Incidence distribution of acute MI type among patients (Descriptive data).

**Figure 2.**
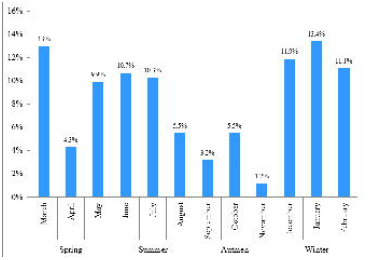
Distribution of monthly and seasonal admission of patients with acute MI (Descriptive data).

### Age

The mean age for AMI patients was 60.4±12.97 years old. There was no significant relationship between the age of the patients with trigger type for AMI (P-value = 0.107).

### Body Mass Index

The mean BMI for the AMI patients were 26.65±4.35 kg/m^2^ (26.40±4.26 for men and 27.40±4.55 for women). There were no significant differences in BMI of the patients with trigger types of AMI (P-value = 0.351).

### Ejection Fraction

The mean left ventricle ejection fraction (LVEF) for the AMI patients was 36±10.4 %. The differences between the mean EF of the AMI patients were significant. So, the EF of the patients with no trigger was significantly lower than patients with physical and mental AMI triggers (P-value = 0.03).

### Gender

In both genders, most patients did not have any triggers in the last 24 hours before AMI (61.9% of women and 50.8% of men respectively). The mental trigger was the most common in both sexes. But, in men, the physical trigger was significantly more than women (P-value = 0.027).

### Residency area

Our results indicated that the type of AMI trigger can relate to the area of residency or living (urban or rural). So, mental triggering in urban living is significantly higher than physical triggers (31.9% vs. 12.9% respectively, P-value = 0.039).

### Occupation

In our study, we assessed the AMI patient occupations to find out whether there is a relationship between job and AMI triggering or not. Generally, in the trigger positive patients, mental triggering was more common than physical triggering. But in retired or housewife patients, physical triggers are much less than mental triggers. The differences between occupations and AMI triggers was significant (P-value = 0.002)

### Diabetes mellitus

In our study 23.4% of patients had diabetes mellitus (18.61% of men and 37.5% of women, P-value = 0.002). Despite gender, our results showed that there are no significant differences between DM and the AMI triggering (P-value = 0.368).

### Hypertension

In AMI patients 41.12% had HTN (36.12% of men and 55.5% of women, P-value = 0.009). There was no significant relationship between trigger types and AMI triggering (P-value = 0.351).

### Smoking

In our study, there was a significant difference in the AMI triggers in smoking and non-smoking patients. So, the physical triggers were more common in smokers than non-smokers (P-value = 0.008).

### Cardiovascular disease history in the family

Most of the patients with and without CVD history in the family didn’t have any trigger for AMI in the past 24h before the attack (43.1% and 56.9% respectively). There was no significant difference between the AMI triggers with cardiovascular disease history in the first-grade family of the patient (P-value = 0.179).

### AMI type

Fig. 1 shows the distribution of AMI type in the patients. Anterior STEMI was the most frequent MI type in patients (45.9%). NSTEMI included 13.4% of all AMIs. In trigger assessment, there were no significant differences in AMI types and AMI triggering (P-value = 0.224).

### Hypercholesterolemia

Over 82% of the AMI patients had hypercholesterolemia (serum total cholesterol ≥130mg/dL). In hypercholesterolemic patients, 26.42% were women and 73.58% were men but, the difference was not significant through gender (P-value = 0.18). Also, the total cholesterol level was not significantly different between the AMI trigger groups (P-value = 0.085).

### High-Density Lipoprotein (HDL)

We assessed the HDL level based on the patient’s gender; So, we determined the “Low HDL” as <50mg/dL for women and <40mg/dL for men. Our results indicated that there were no significant differences in low levels of serum HDL with the type of AMI triggers (P-value = 0.217).

### Low-Density Lipoprotein (LDL)

In the analysis of serum LDL levels of the patients, our data indicated that there were no significant differences in serum LDL levels (two groups of normal LDL and high LDL) with the type of AMI triggers (P-value = 0.056).

### Serum Triglyceride

Most of the patients with and without abnormal TG didn’t have any activity in the past 24 hours before AMI. We assessed serum TG two times; first, we grouped patients to normal TG, borderline TG, or high TG levels. The difference between TG groups and AMI triggers was significant (P-value = 0.035). On the second time, we regrouped patients to normal TG and abnormal TG (borderline and high). At this time also, the difference between TG groups and AMI triggers was significant (P-value = 0.018).

### Season

Considering the one-year study, we assessed the differences in AMI triggers with differences in the seasons. Our results indicated that the difference between a season of AMI occurrence and AMI triggers was significant and the physical triggers decreased in autumn and winter (P-value = 0.013). There were two peaks of AMI incidence regardless of the triggers; the First one was from December to March, and the second one was from May to July (Fig. 2).

## Discussion

In the 21^st^ century, despite the progression of medical sciences and technology advances, cardiovascular diseases remain major public health problem in different countries, so that caused 31% of global deaths in 2015 and 7.4 million of these deaths were due to coronary heart disease [2, 14]. Myocardial infarction is a type of coronary heart disease that is defined as necrosis of the heart muscle resulting from ischemia or supply-demand imbalance [15]. There are some definite risk factors such as diabetes, hypertension, abnormal obesity, impaired lipid profile, smoking, and psychological stresses [16-21]. But still, the one question remains unanswered. Are there some factors that may induce myocardial infarction in individuals with or without risk factors? If yes, what are they and how important they are? Some studies have been done to answer the questions. Andrew Smyth and et al, have reported that physical exertion and anger or emotional upset can be external triggers for acute myocardial infarction in the next 1 hour [22]. In other study, Jeremy Ben-Shoshan et al have reported MI trigger in 37% of patients with acute MI with majority of physical activity (67%) as potential trigger [23]. But also there are studies that reported up to half ratio for triggering or prodromal factors in acute MI [24].

Our study revealed 53% of patients with acute MI, had no kind of defined trigger in the past 24 hours before MI symptoms. Despite the mentioned studies that have evaluated single or a couple of prodromal factors, in this study, we assessed all possible and accessible triggering factors candidates, so that the other 43% had a kind of triggering activity including heavy activity (physical), insomnia or mental tensions (for a while before symptoms), overeating or noisy parties, receiving bad news, anger or fighting, overdosing of drugs or opiates to frequency but due to the small number of patients in most of the subgroups we regrouped them as three main group of the Physical triggers, the Mental triggers, and No triggers. The difference between the two sexes was significant revealing triggers are effective in males susceptible to acute MI three times more than females but there were no significant differences for age groups. Monthly and seasonal admission reveals two peaks, the first one in winter and related to cold weather that is confirmed by previous studies [25, 26], and the second one was in summer. According to sex and occupational distribution and the significance of trigger induction in physical triggers related jobs; we hypothesize that the summer peak maybe is related to an increase in physical activity due to daylight increase and farming activity. Also, this hypothesis is confirmed by a sudden decrease after March; Iranian new year holidays or “Nowrooz” (7 to 15 days) beginning after March 20th same to past studies [24]. By the way, living area differences revealed a significant relationship that can be related to civic and urban living daily stresses. Hypertriglyceridemia is a major risk factor for cardiovascular disease and in our study, it has been demonstrated that despite hypercholesterolemia, patients with hypercholesterolemia have a significant risk for MI induced by prodromal triggers. According to our findings, in patients with a history of diabetes, the mentioned triggers would not induce acute MI. In diabetic patients, this may relate to autonomic neuropathy [27, 28] but we cannot be certain about that. In this study, we couldn’t find a significant relationship between age, BMI, diabetes mellitus, hypertension, familial history of cardiovascular diseases, AMI type, hypercholesterolemia, low HDL levels, and high LDL levels but the results revealed significant differences in AMI triggers with ejection fraction (EF), gender, residency and living area, occupation, smoking, abnormal serum TG levels and the season of AMI occurrence.

In this study, we concluded that in addition to classic risk factors of ischemic heart disease and myocardial infarction, health care systems and physicians must pay more attention to triggers that may induce an acute myocardial infarction in people with predisposing factors specially in male sex, stressful and hand working jobs, and psychological and mental tension patients.

## Data Availability

Not available for public

## Acknowledgements

We would like to thank the health care personnel of Seyedoshohada heart center, specially the angiography and echocardiography ward personnel.

## Clinical Significance

- Physical and mental triggers can be found in almost half of acute myocardial infarctions.
- Health care systems can decrease a great fraction of risk for acute myocardial infarctions, by educations and health care preventions of these triggers.

## Contributorship Statement

In this study all authors have contributed equally in idea development, design, data collection, study performance, writing the draft and review.

## Patient and Public Involvement

The study was supported by a patient advisory group which provided input to the program of research. This patient advisory group met on a regular basis for the duration of the study. Patients partnered with us for the design of the study, the informational material to support the intervention, and the burden of the intervention from the patient’s perspective. At the end of the study, the patient advisory group commented on the findings and contributed to the dissemination plan.

## Text Table

**Table 1.** Clinical and paraclinical data of the study with analysis for acute MI triggers. *The reported values are mean ± standard deviation and P-values are estimated based on one-way ANOVA. The reported values for other variables in the table are number (%), and P values are estimated based on the chi-square test.

